# Ultra-Sensitive High-Resolution Profiling of Anti-SARS-CoV-2 Antibodies for Detecting Early Seroconversion in COVID-19 Patients

**DOI:** 10.1101/2020.04.28.20083691

**Authors:** Maia Norman, Tal Gilboa, Alana F. Ogata, Adam M. Maley, Limor Cohen, Yongfei Cai, Jun Zhang, Jared E. Feldman, Blake M. Hauser, Timothy M. Caradonna, Bing Chen, Aaron G. Schmidt, Galit Alter, Richelle C. Charles, Edward T. Ryan, David R. Walt

**Affiliations:** Department of Pathology, Brigham and Women’s Hospital, Boston, MA; Wyss Institute for Biologically Inspired Engineering at Harvard University, Boston, MA; Tufts University School of Medicine, Boston, MA; Harvard Medical School, Boston, MA; Department of Chemical Biology, Harvard University, Boston, MA; Division of Molecular Medicine, Boston Children’s Hospital, Boston, MA; Ragon Institute of MGH, MIT and Harvard, Cambridge, MA; Department of Pediatrics, Harvard Medical School, Boston, MA; Department of Microbiology, Harvard Medical School, Boston, MA; Division of Infection Diseases, Massachusetts General Hospital, Boston, MA; Department of Medicine, Harvard Medical School, Boston, MA; Department of Immunology and Infectious Diseases, Harvard T.H. Chan School of Public Health, Boston, MA

## Abstract

The COVID-19 pandemic continues to infect millions of people worldwide. In order to curb its spread and reduce morbidity and mortality, it is essential to develop sensitive and quantitative methods that identify infected individuals and enable accurate population-wide screening of both past and present infection. Here we show that Single Molecule Array assays detect seroconversion in COVID-19 patients as soon as one day after symptom onset using less than a microliter of blood. This multiplexed assay format allows us to quantitate IgG, IgM and IgA immunoglobulins against four SARS-CoV-2 targets, thereby interrogating 12 antibody isotype-viral protein interactions to give a high resolution profile of the immune response. Using a cohort of samples collected prior to the outbreak as well as samples collected during the pandemic, we demonstrate a sensitivity of 86% and a specificity of 100% during the first week of infection, and 100% sensitivity and specificity thereafter. This assay should become the gold standard for COVID19 serological profiling and will be a valuable tool for answering important questions about the heterogeneity of clinical presentation seen in the ongoing pandemic.

## Main

SARS-CoV-2 (severe acute respiratory syndrome coronavirus-2) is a novel beta- coronavirus that has caused the coronavirus disease pandemic of 2019 (COVID-19) ^1–3^. In order to identify infected individuals and contain the spread of the disease, rapid and accurate population-wide screening is essential. Currently, nucleic acid based tests of nasopharyngeal (NP) swabs are the primary method used to detect COVID-19 infected individuals ^2^. These tests can only diagnose disease during a narrow window of active infection with an overall clinical sensitivity of 65-72% ^4^. Thus, additional diagnostic methods are needed to identify those who are or have been infected with SARS-CoV-2. Robust serological assays that can detect antibodies against SARS-CoV-2 can fulfill this critical need. These assays could be used to detect infected individuals who have tested negative by RT-PCR. Furthermore, serological assays could be used to identify individuals who have been infected with SARS-CoV-2 and were asymptomatic or had mild symptoms, thereby providing a better understanding of how widespread the virus is within a population. This information can aid epidemiologists in determining a more accurate population prevalence of COVID-19.

Many serological enzyme-linked immunosorbent assays (ELISA) have been recently developed to detect anti-SARS-CoV-2 antibodies. However, these assays have four important limitations ^4–15^. First, they lack the ability to detect antibodies at early stages of infection. Second, false positive results due to non-specific binding can occur attributable to high levels of pre-existing antibodies in blood ^16^. Third, conventional ELISA assays can only analyze one immunoglobulin for a specific target at a time, limiting their ability to profile and understand the underlying immune response. Finally, these assays lack the resolution to quantify variations in the immune response, which may be key in understanding clinical progression.

To address these limitations, we developed ultra-sensitive Single Molecule Array (Simoa) assays for anti-SARS-CoV-2 IgG, IgM, and IgA antibodies against four immunogenic viral proteins, providing us with detailed information about early stages of immune activation ^17^. Simoa provides up to 1,000-fold improvement in analytical sensitivity over standard ELISA ^18,19^. This ultra-sensitivity enables samples to be highly diluted resulting in significantly reduced non-specific binding. Additionally, unlike the standard ELISA, Simoa has a wide dynamic range and allows precise quantification of multiple analytes over a concentration range of four orders of magnitude. This feature is particularly important since antibody levels vary between different stages of infection, and thus it is advantageous to have a single assay format that can quantitatively differentiate between individuals with high and low antibody levels.

We measured the levels of all three immunoglobulin isotypes binding to the four viral proteins in patients who were determined COVID-19 negative or positive by NP RT-PCR testing. We also measured the immunoglobulins in plasma samples collected from patients before the start of the COVID-19 pandemic. By looking at two time points for each positive NP RT-PCR patient, we were able to study the response of the immune system over time. We show that these serological assays detect anti-SARS-CoV-2 antibodies with high sensitivity and specificity at both earlier and later stages of infection.

### Developing an ultra-sensitive Simoa assay for anti-SARS-CoV-2 antibodies

We developed a multiplexed ultra-sensitive Simoa assay for detection of IgG, IgM, and IgA against SARS-CoV-2 in human plasma. This multiplexed assay detects the binding of each of the three immunoglobulin isotypes to four viral targets: Spike Protein, S1 subunit, Receptor Binding Domain (RBD), and Nucleocapsid, enabling the quantification of 12 binding interactions. In this assay format (Figure 1), four types of dye-encoded paramagnetic beads are each coated with one of the four SARS-CoV-2 targets. The coated beads are then incubated with diluted heat-inactivated plasma samples for capture of immunoglobulins on the beads. A large excess of beads is used compared to the number of immunoglobulin molecules in the sample, such that either zero or one immunoglobulin molecule binds to each bead. After immunoglobulin capture, the beads are incubated with a biotinylated anti-human immunoglobulin antibody specific to a single immunoglobulin isotype. Finally, the beads are incubated with the enzyme streptavidin-β-galactosidase (SβG), which binds to the biotinylated anti-human immunoglobulin antibody, forming a complete enzyme-labeled immunocomplex. The beads are then resuspended in resorufin β-D-galactopyranoside (RGP), a fluorogenic substrate, and loaded onto an array of femtoliter-sized wells in which only one bead fits per well. The wells are sealed with oil and a locally high concentration of the fluorescent product is generated. This enables single molecule detection of immunoglobulins by counting active wells. The ratio of active wells to the total number of beads is determined for each target, and the Average Enzymes per Bead (AEB) is calculated, providing a digital readout for each immunoglobulin isotype against each antigen.

**Figure 1.**
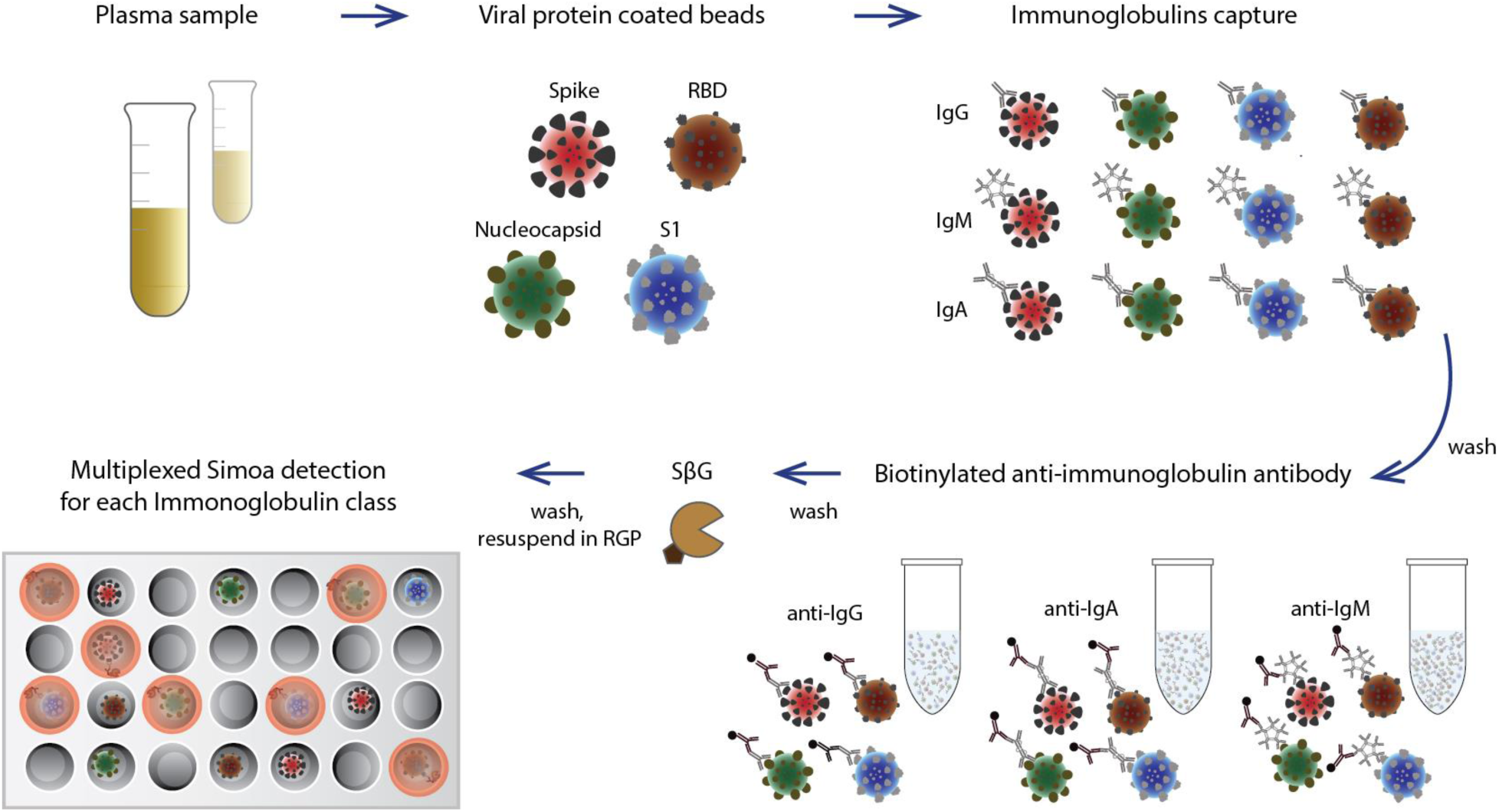
Schematic illustration of the Simoa serological assay. Inactivated plasma is incubated with four types of dye-encoded beads that are each coupled to one of four viral antigens (Spike, S1, Receptor Binding Domain, and Nucleocapsid). IgG, IgA, and IgM antibodies specific to the SARS-CoV-2 antigens bind to the viral antigen coated beads. After washing, beads are introduced to biotinylated anti-human immunoglobulin antibodies to label either IgG, IgM, or IgA in the different reactions. After additional washes, the enzyme streptavidin-β-galactosidase (SβG), is introduced. The beads are washed, resuspended in fluorogenic resorufin β-D-galactopyranoside (RGP) and loaded into a 216,000 microwell array for multicolor imaging.

During the assay development process, we ensured optimal performance of the serological Simoa assays at several different steps. We produced and purified the SARS-CoV-2 Spike protein and the smaller RBD (Methods section and Supplementary Figures 1 and 2), while the Nucleocapsid and S1 proteins were obtained commercially. We quantitatively validated viral target conjugation to the bead surface using anti-His tag antibodies as well as recombinant human anti-RBD antibody, as described in the Supplementary Information (Supplementary Figure 3). The detector antibody for each immunoglobulin isotype was selected from a panel of antibodies to optimize the performance of the assay (Supplementary Tables 1–3). Finally, we performed spike-and-recovery and dilution-linearity experiments using recombinant human immunoglobulins to demonstrate assay precision and validity. (Supplementary Figure 4 and Supplementary Table 4). The Simoa assays show excellent linearity and recoveries ranging from 85–103%.

**Figure 2.**
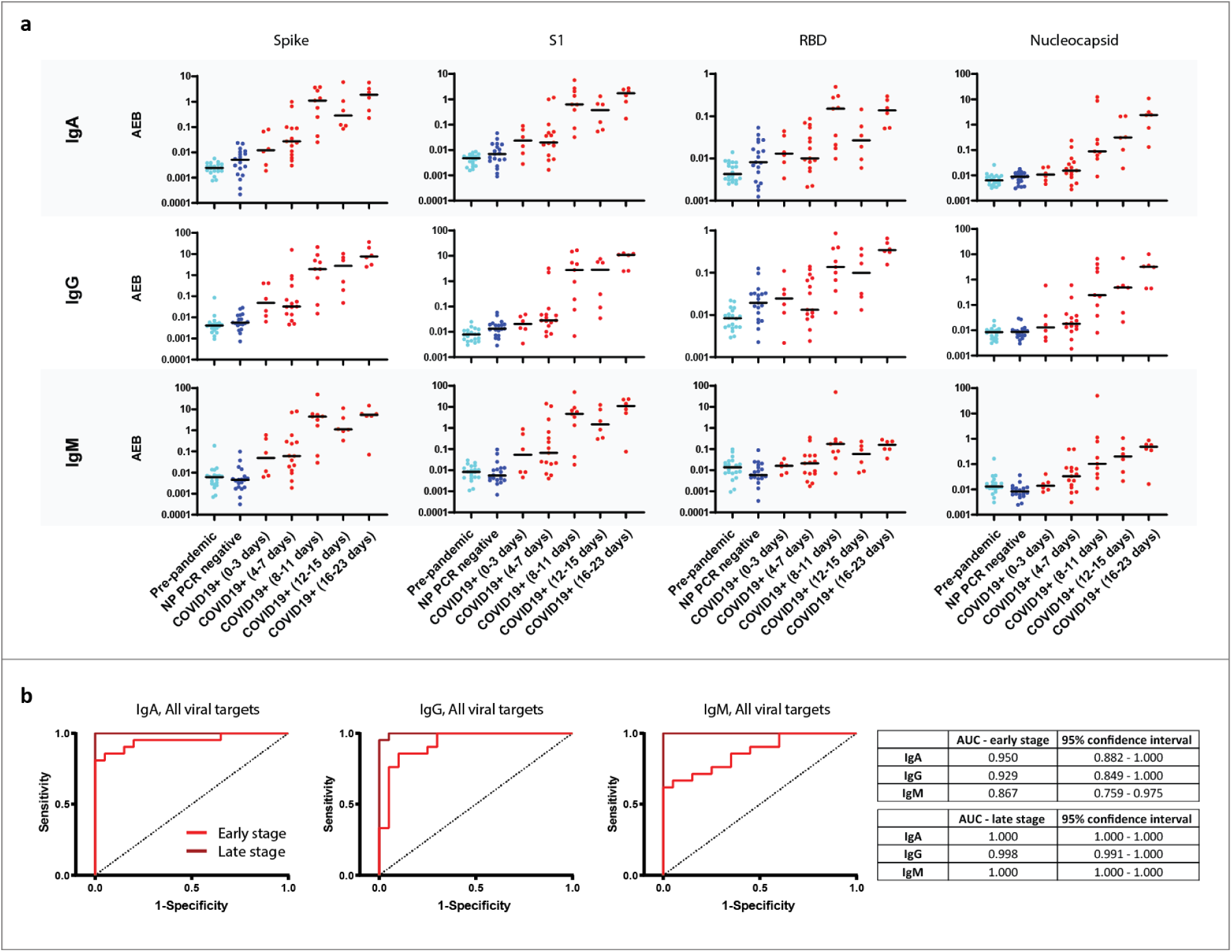
Profiling seroconversion in COVID-19. a) Simoa serological assay results for IgG, IgM, and IgA against the four viral targets: Spike, S1 subunit, Receptor Binding Domain, and Nucleocapsid for pre-pandemic samples (light blue, n=20), NP PCR negative samples (dark blue, n= 19) and COVID-19 positive samples (red, n=21, two timepoints each). The COVID-19 positive samples were divided into five groups according to time since symptom onset. Black lines indicate the mean AEB (average enzyme per bead) value of each population. b) Receiver Operating Characteristic (ROC) curves for all four antigens against IgA, IgG, and IgM. Each ROC graph is plotted for COVID-19 disease within one week of symptom onset (red, n=21), and for later stage COVID-19 (maroon, n=21) each against the pre-pandemic cohort (left, n=20). AUC values and 95% confidence interval are presented in the table (right).

### Detection of anti-SARS-CoV-2 antibodies in patient samples using Simoa

We used the Simoa serological assay to test 81 plasma samples from Massachusetts General Hospital (MGH) in Boston, MA, USA. This sample cohort consisted of three groups: (1) patient plasma collected before the COVID-19 outbreak, (2) plasma collected during the pandemic from patients presenting with symptoms of SARS-CoV-2 at the Emergency Department (ED) with a negative NP RNA test, and (3) serial plasma samples collected during the pandemic from patients who had tested positive for SARS-CoV-2 infection and were hospitalized for COVID-19. Clinical characteristics of each patient group are shown in Supplementary Table 5. Figure 2A shows IgG, IgM, and IgA levels in COVID-19 positive samples, which were divided into five subgroups based on the number of days after symptom onset. In addition, Figure 2A shows the immunoglobulin levels in the pre-pandemic group and the NP RT-PCR negative group. Samples were measured at two different dilution factors and similar results were observed (Supplementary Figure 5). Plasma samples measured at a 4000x dilution factor show that all four viral antigens elicit an IgG, IgM, and IgA immune response in COVID-19 positive patients. All twelve combinations (i.e. each of the four viral targets against each of the three immunoglobulins) show an overall increase in immunoglobulin levels over the course of the disease. Only one patient had a decrease in immunoglobulin levels; this person did not survive their infection with SARS-CoV-2. In addition, a separate patient who displayed low immunoglobulins levels at the late time point was on immunosuppressive medication.

To study the ability of the serological Simoa assays to detect seroconversion early relative to symptom onset, we separated the results of the COVID-19 positive samples into two sub-groups. The first group consists of data from early time points, in which samples were collected in the first week following symptom onset. The second group consists of data from late time points, in which samples were collected after the first week of symptom onset. We assessed the performance of the IgG, IgM, and IgA Simoa assays using multivariate analysis for each of the early and late time point groups compared to the pre-pandemic group. Figure 2B shows the receiver operating characteristic (ROC) curves for IgG, IgM, and IgA against all four viral targets. For the early time point group, IgA had the highest area under the curve (AUC) at 0.95 followed by IgG at 0.93 and IgM at 0.87. The sensitivity of the IgA assay was 81% during the first week at 100% specificity. Moreover, the AUC values for all three immunoglobulins improve for the late stage group with IgA at 1, IgG at 1, and IgM at 0.998.

Next, we plotted the ROC curves for each of the 12 viral target and immunoglobulin combinations to understand which viral antigens are critical to measuring the immune response by classifying between COVID-19 positive groups and pre-pandemic groups (Supplementary Figure 6). The Spike protein gave the highest sensitivity and specificity for each immunoglobulin subtype at early time points, with a calculated AUC of 0.94, 0.93 and 0.85 for IgA, IgG and IgM respectively. The Spike IgA assay yielded the best classification when comparing the pre-pandemic group to the COVID-19 positive group for early time points.

A major advantage of the Simoa serological assay is that one sample provides information on all three immunoglobulin responses to four viral proteins compared to a traditional ELISA where only a single interaction can be interrogated. Therefore, based on our analysis in Supplementary Figure 6, we sought to assess the sensitivity and specificity of a combination of IgA and IgG assays against spike protein for early time points. We performed a multivariate analysis using only the combination of IgA and IgG Spike assays and were able to achieve a sensitivity of 86% and a specificity of 100% for identifying patients within the first week of symptom onset and 100% sensitivity and specificity for identifying patients after the first week of symptom onset (Figure 3). We also assessed the positive predictive values (PPV) and negative predictive values (NPV) of the combined Spike IgA/IgG assays for the two groups (Figure 3B). As the true prevalence of COVID-19 in the population is unknown, varies among countries, and is rapidly changing, we calculated these values for a range of prevalence values (Figure 3B). We also sought to evaluate whether patients who tested negative for COVID-19 had antibodies against SARS-CoV-2 viral antigens in their blood. These individuals were tested by NP RT-PCR since there was clinical suspicion that they may have been infected with the virus. Interestingly, when assessing this RT-PCR COVID-19 negative group, 9 of the 19 patients were above the seroconversion threshold. This result is in line with the RT-PCR tests having a reported clinical sensitivity between 65-72% ^4^. It is possible that these patients with negative RNA test results were infected with SARS-CoV-2 and were seroconverting.

**Figure 3.**
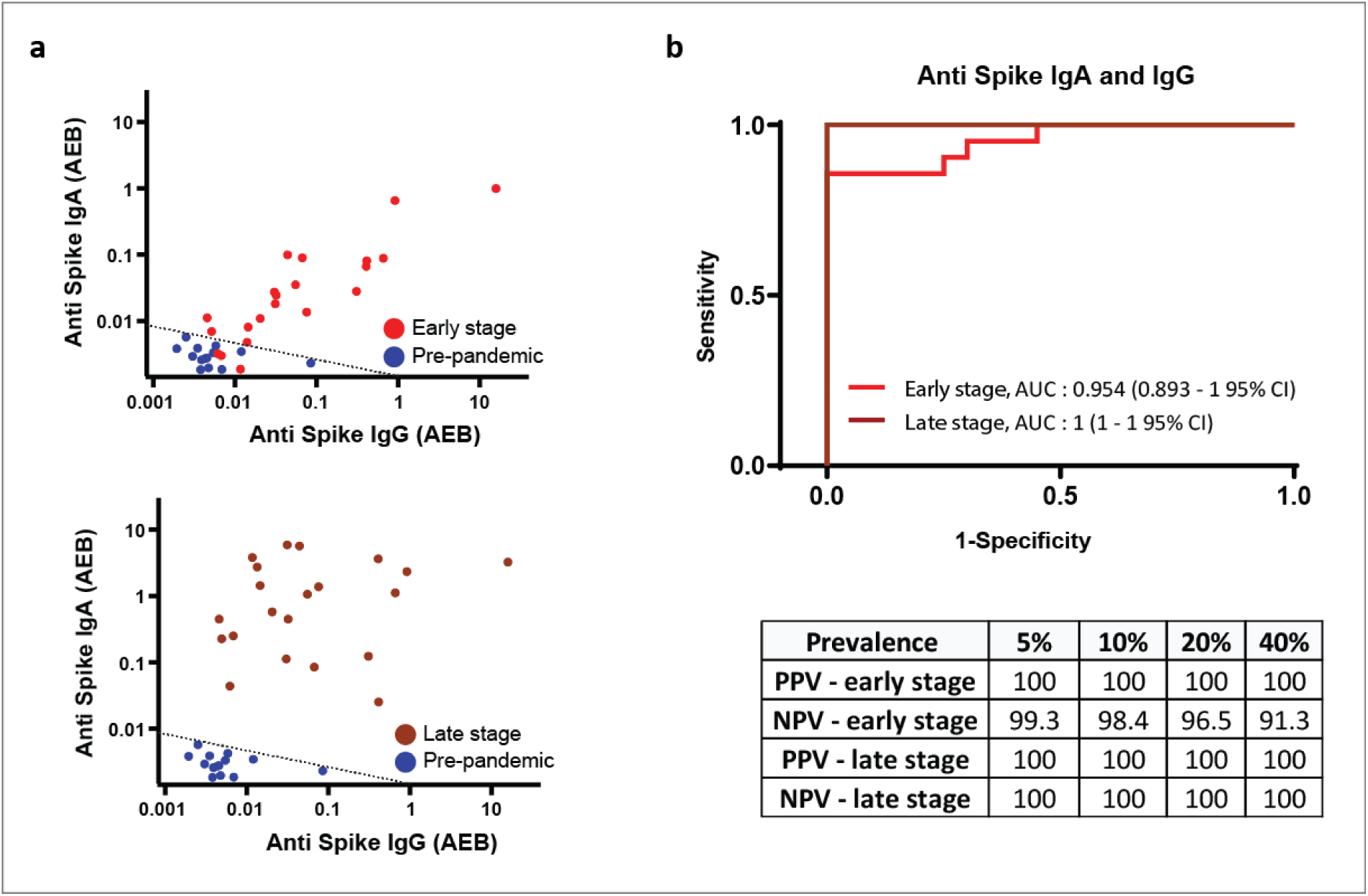
Classification of COVID-19 disease using Simoa serological assays. a) Scatterplots of the anti-spike IgG and IgA immunoglobulins for COVID-19 disease within one week of symptom onset (top, red, n=21), and for later stage COVID-19 (bottom, maroon, n=21) each against the pre-pandemic cohort (blue, n=20). Dotted black lines represent the classification cutoff. b) Multivariant Receiver Operating Characteristic (ROC) curve and AUC values for anti-spike IgG and IgA immunoglobulins plotted for COVID-19 disease within one week of symptom onset (red, n=21), and for later stage COVID-19 (maroon, n=21) each against the pre-pandemic cohort (blue, n=20). A table of positive and negative predictive values for a range of prevalence data is displayed (bottom).

## Discussion

Quantitative and sensitive SARS-CoV-2 serological assays are necessary to address unmet needs in the clinical setting, epidemiological studies, as well as therapeutic and vaccine development. Using the Simoa technology, we developed a highly sensitive serological assay for detection of IgG, IgM, and IgA in COVID-19 patients against four SARS-CoV-2 targets. These assays were optimized for use on an automated commercially-available instrument, which enables rapid, reproducible and high throughput results. The ultra-sensitivity enables plasma to be diluted 4000x allowing for improved classification of COVID19 patients, likely due to negligible nonspecific binding. Furthermore, at this dilution factor our dynamic range spans four orders of magnitude, allowing for quantification of the full range of disease from early to late stage. This dilution factor requires a sample volume of less than 1μL plasma, potentially enabling wide scale epidemiological testing with a finger prick sample collection method or dried blood spots.

Since the clinical sensitivity of current RT-PCR tests is approximately 70%, it was crucial to establish a true negative sample group in addition to the negative NP RNA test group. Our analysis includes samples collected from U.S. adults presenting to the MGH Travel and Immunization Center prior to traveling internationally. These samples were collected January-December 2019 before introduction of SARS-CoV-2 into the U.S. Although we cannot comment on active non-SARS-CoV-2 coronoviridae infection nor previous exposure to such viruses among these individuals, such viruses are a common cause of respiratory tract infections. Our results suggest no significant cross reactivity of antibodies to our selected target antigens prior to the COVID-19 outbreak, indicating minimal cross reactivity among coronaviridae in our assay.

In our dataset, IgA and IgG showed superior performance for early seroconversion detection. However, the samples we used were heat inactivated, which has been shown to be detrimental for serological assays and particularly the ability to detect IgM antibodies ^20^. It will be important to investigate whether using non-heat inactivated plasma or serum can yield higher sensitivity for the IgM assay, potentially leading to improved acute stage diagnosis. In addition, since our assay provides detailed and high-resolution information about the immune response to different SARS-CoV-2 antigens, the assay format should enable us to predict neutralization potential of convalescent plasma.

Using a combination of IgG and IgA responses to spike protein, we are able to achieve a sensitivity of 86% and a specificity of 100% during the first week of infection when comparing COVID-19 positive patients to pre-pandemic controls. This sensitivity increased to 100% and the specificity remained at 100% for COVID-19 positive patients tested one week after symptom onset. This early detection of seroconversion is unmatched by current commercial tests or published laboratory data. Additionally, the resolution of our method enables us to probe small variations in the immune response between patients and within a single patient. The Simoa serological platform provides a powerful analytical tool that will benefit future research in understanding the immune response to COVID-19 by enabling the analysis of antibody response at the early stages of the disease and throughout disease progression with high resolution. A comprehensive analysis of the immune response can provide critical insights necessary for the development of therapeutics and vaccines.

## Methods

### Plasma samples

Clinical samples were obtained from patient presenting to the Massachusetts General Hospital with viral respiratory symptoms. COVID-19 infection was confirmed by RT-PCR on nasopharyngeal specimen. Additional plasma samples collected from healthy adults pre-pandemic (January–December 2019) prior to travel internationally were also included in the analysis. All samples were collected under approval of the Institutional Review Board for Human Subjects Research at Massachusetts General Hospital. Human plasma samples were heat inactivated (56°C for 60 minutes). Samples were hand diluted in sample buffer to their final dilution factor which varied for each experiment, as described in the results section. All sample measurements had two technical replicates which were averaged.

### RBD Expression and Purification

SARS-2 RBD (GenBank: MN975262.1) was cloned into pVRC vector for mammalian expression (FreeStyle 293F or Expi293F suspension cells). The construct contains a HRV 3C-cleavable C-terminal SBP-His_8X_ tag. Supernatants were harvested 5 days post-transfection and passaged directly over Cobalt-TALON resin (Takara) followed by size exclusion chromatography on Superdex 200 Increase (GE Healthcare) in 1x phosphate-buffered saline. Typical yields from FreeStyle 293F cells are approximately 9 mg/liter culture. Affinity tags were removed using HRV 3C protease (ThermoScientific) and the protein repurified using Cobalt-TALON resin to remove the protease, tag and non-cleaved protein. RBD sequence and purification validation are provided in the Supplementary Figure 1.

### Preparation of a stabilized ectodomain of Spike protein

To express a stabilized ectodomain of Spike protein, a synthetic gene encoding residues 1–1208 of SARS-CoV-2 Spike with the furin cleavage site (residues 682–685) replaced by a “GGSG” sequence, proline substitutions at residues 986 and 987, and a foldon trimerization motif followed by a C-terminal 6xHisTag was created and cloned into the mammalian expression vector pCMV-IRES-puro (Codex BioSolutions, Inc, Gaithersburg, MD). The expression construct was transiently transfected in HEK 293T cells using polyethylenimine. Protein was purified from cell supernatants using Ni-NTA resin (Qiagen), the eluted fractions containing S protein were pooled, concentrated, and further purified by gel filtration chromatography on a Superose 6 column (GE Healthcare), following a protocol described previously ^21^. Negative stain electron microscopy (EM) analysis was performed as described ^22^. Additional information is provided in Supplementary Figure 2

### Bead Coupling & Verification

Recombinant SARS-CoV-2 antigens (Nucleocapsid, RBD, S1 and Spike) were coupled to four types of dye-encoded 2.7 μm carboxylated paramagnetic beads (Quanterix) using EDC (1-ethyl-3-(3-dimethylaminopropyl)carbodiimide hydrochloride) chemistry (ThermoFisher Scientific 77149). The Nucleocapsid and S1 antigens were sourced commercially (Nucleocapsid: Ray Biotech 230-30164 and S1: Sino Biological V0591-V08H). The Spike and RBD antigens were produced in the labs of Bing Chen and Aaron Schmidt, respectively, as described above. 2.8 × 10^8^ beads were washed three times with 200 μL of Bead Wash Buffer (Quanterix), three times 200 μL of Bead Conjugation Buffer (Quanterix), and then resuspended in 300 μL of Bead Conjugation Buffer. Immediately prior to use, 10 mg of EDC was reconstituted in 1 mL of Bead Conjugation Buffer. To activate the beads for conjugation, EDC was added to the bead suspension and the beads were agitated on a HulaMixer (ThermoFisher Scientific) for 30 minutes at 4°C. After activation, the beads were washed once with 200 μL of Bead Conjugation Buffer, and then resuspended in 300 μL of Bead conjugation buffer containing the antigen. Beads were agitated on the HulaMixer for 2 hours at 4°C. The antigen-conjugated beads were washed two times with 200 μL of Bead Wash Buffer, and then blocked with BSA (Bovine serum albumin) for 30 minutes at room temperature in 200 μL of Bead Blocking Buffer (Quanterix). The blocked beads were washed with 200 μL of Bead Wash Buffer, 200 μL of Bead Diluent (Quanterix), and resuspended in 200 μL of Bead Diluent. The amount of recombinant protein and EDC used in each reaction are as follows: Nucleocapsid (20μg antigen, 2μL EDC), RBD (20μg antigen, 6μL EDC), S1 (20μg antigen, 6μL EDC), and Spike (16.67μg antigen, 6μL EDC). Nucleocapsid, Spike, S1, and RBD were conjugated to 488 nm, 647 nm, 700 nm, and 750nm dye-encoded beads, respectively. Beads were counted using a Beckman Coulter Particle Counter and stored at 4°C.

Antigen coupling to the beads was confirmed by an anti-His tag assay for Spike, S1, and nucleocapsid beads and by an anti-RBD assay for RBD beads (Supplementary figure 3). For spike, S1, and nucleocapsid, confirmation of antigen attachment to the beads was demonstrated by Simoa with His tags experiments using a biotinylated anti-His tag antibody (ThermoFisher MA121315BTI) on the HD-X Analyzer (Quanterix). The anti-His-tag antibody was plated at concentrations of 0.1 pg/mL to 10,000 pg/mL using tenfold dilutions. RBD was provided without a His-tag, therefore RBD beads were used as a control against the anti-His tag antibody assay. RBD conjugation to beads was confirmed by Simoa with an anti-RBD antibody (clone CR3022) and a biotinylated anti human-IgG antibody (Bethyl Laboratories A80–148B). Nucleocapsid beads were used as a control for these experiments due to the ability of anti-RBD antibody binding to RBD, S1, and full Spike protein but not nucleocapsid.

### Biotinylation

Detection antibodies for IgA, IgG and IgM were purchased from Thermo Fisher, Bethyl Laboratories, Abcam, Biolegend, and R&D systems (see Immunoglobulin Simoa assay format) and were biotinylated for use in Simoa assays as described previously by Cohen et al ^23^. Briefly, the antibodies were passed through an Amicon filter three times in Biotinylation Reaction Buffer (Quanterix). Antibody concentrations were determined using NanoDrop One Spectrophotometer. Antibodies were conjugated to biotin using EZ-Link NHS-PEG4 Biotin (Thermo Fisher Scientific) by resuspending NHS-PEG4-Biotin in dionized H_2_O. For all immunoglobulins a NHS-PEG4-Biotin was added in 40x molar excess and incubated for 30 min. All biotinylated antibodies were then purified using three washes in an Amicon filter.

### Immunoglobulin Simoa assay format

Simoa experiments were performed in an automated three-step assay format onboard the HD-X Analyzer (Quanterix Corp) as described in Rivnak et al ^24^. Human plasma samples were diluted in Homebrew Detector/Sample Diluent (Quanterix). Anti-human immunoglobulin antibodies were diluted in Homebrew Detector/Sample Diluent to final concentrations of: IgG (Bethyl Labratories A80–148B): 7.73ng/mL, IgM (Thermo Fisher MII0401): 216ng/mL, IgA (Abcam ab214003): 150ng/mL. Streptavidin-β-galactosidase (SβG) concentrate (Quanterix) was diluted to 30 pM in SβG Diluent (Quanterix). System Wash Buffer 1, System Wash Buffer 2, Resorufin β-D-galactopyranoside (RGP), and Simoa Sealing Oil were purchased from Quanterix and loaded onto the HD-X Analyzer per the manufacturer’s instructions. In the first step of the assay, 25μL of the four SARS-CoV-2 antigen-coupled multiplex beads were incubated with 100μL of diluted human plasma for 15 minutes. The total number of beads used per reaction was 475,000 (125,000 each of Nucleocapsid, RBD, and S1 beads and 100,000 of Spike beads). After incubation, six wash steps were performed with System Wash Buffer 1. In the second step, the beads were resuspended in 100 μL of the respective biotinylated anti-human immunoglobulin antibody and incubated for 5.25 minutes, and then washed six times with System Wash Buffer 1. In the third step, the beads were resuspended in 100μL of SβG, incubated for 5.25 minutes and washed six times. The beads were resuspended in 25 μL of

RGP before being loaded into the microwell array for analysis. Following bead loading, the microwell array was sealed with oil and imaged in five optical channels. Average Enzyme per Bead (AEB) values were calculated by the software in the HD-X Analyzer.

## Data Analysis

Duplicate measurements per sample were obtained for each of the twelve immunoglobulin and viral target combinations. The average of duplicate measurements was calculated and the data was then log transformed. Logistic regression analysis was conducted in R version 3.6.2 for the multivariate analysis and Graphpad Prism 7 for the univariate analysis^25^. All figures were plotted in Graphpad Prsim 7, Igor Pro7 and Adobe Illustrator version 2015.

## Data Availability

The authors declare that the data supporting the findings of this study are available within the paper and its Supplementary information files.

## Acknowledgments

The authors would like to thank Liangxia Xie for the helpful discussions regarding the experimental design. This work was largely funded through a grant from the Massachusetts Consortium for Pathogen Resilience. In addition, support came from the Global TravEpiNet (GTEN) system sponsored by the US Centers for Disease Control and Prevention (Grant No. U01CK000490: ETR, RCC) as well as a T32GM007753 grant from NIGMS, a T32 AI007245 from NIAID, and an R01AI146779 from NIAID.

## Author Contributions

M.N., T.G., A.F.O., A.M.M., L.C., and D.R.W. conceived the approach. M.N., T.G., A.F.O. and A.M.M. performed the experiments, M.N., T.G., and L.C. analyzed the data, Y.C., J.Z., J.E.F., B.M.H., T.M.C., B.C., and A.G.S. produced and purified the antigens, R.C.C. and E.T.R. collected the samples, M.N., T.G., A.F.O., L.C., A.M.M. and D.R.W. co-wrote the paper. All authors were involved in designing experiments, reviewing and discussing data, and commented on the manuscript.

## Competing interests

David Walt has a financial interest in Quanterix Corporation, a company that develops an ultra-sensitive digital immunoassay platform. He is an inventor of the Simoa technology, a founder of the company and also serves on its Board of Directors. Dr. Walt’s interests were reviewed and are managed by BWH and Partners HealthCare in accordance with their conflict of interest policies.

The assays in this publication have been licensed by Brigham and Women’s Hospital to Quanterix Corporation.

## Supplementary Information is available for this paper

**Data Availability Statement:** The authors declare that the data supporting the findings of this study are available within the paper and its Supplementary information files.

